# Parental education and occupation in relation to risk of childhood type 1 diabetes: nationwide cohort study

**DOI:** 10.1101/2022.07.26.22277835

**Authors:** Paz Lopez-Doriga Ruiz, German Tapia, Inger J. Bakken, Siri E. Håberg, Hanne L. Gulseth, Torild Skrivarhaug, Geir Joner, Lars C. Stene

**Affiliations:** Department of Chronic Diseases, Norwegian Institute of Public Health, Oslo, Norway; Department of Endocrinology, Morbid Obesity and Preventive Medicine, Oslo University Hospital, Oslo, Norway; Norwegian Directorate of Health, Trondheim, Norway; Centre for Fertility and Health, Norwegian Institute of Public Health, Oslo; Institute of Clinical Medicine, University of Oslo, Oslo, Norway; Division of Pediatrics and Adolescent Medicine and Oslo Diabetes Research Centre, Oslo University Hospital, Oslo, Norway

## Abstract

**Background:** Socioeconomic status in the risk of developing type 1 diabetes seems inconsistent. We investigated whether risk of childhood-onset type 1 diabetes differed by parental education or occupation in a nationwide cohort.

**Methods:** This cohort study included all children born in Norway from 1974 to 2013. In individually linked data from nationwide population registries following children born in Norway up to 15 years of age, we identified 4647 with newly diagnosed type 1 diabetes during 15,381,923 person-years of follow-up.

**Results:** Children of mothers with a master’s degree had lower risk of type 1 diabetes than children of mothers with completed upper secondary education only: adjusted incidence rate ratio, aIRR=0.81 95% confidence interval: 0.69 - 0.95). There was no difference between upper secondary and lower secondary maternal education (aIRR=0.98, 95% confidence interval 0.89-1.08). Paternal education was not significantly associated with type 1 diabetes. While maternal elementary occupation was associated with a lower risk of type 1 diabetes, specific maternal- or paternal occupations were not.

**Conclusions:** Our results suggested inverse U-shaped associations between maternal socioeconomic status and risk of type 1 diabetes. Non-linear associations be part of the reason why previous literature has been inconsistent.

## Background

Type 1 diabetes is among the most common chronic diseases in children, and is associated with long-term severe complications and increased mortality [1]. The incidence of childhood-onset type 1 diabetes is generally higher in wealthier countries [2]. The increasing incidence over the past few decades strongly imply early life environmental factors in the aetiology, but few such factors have been identified [3-5].

Many of the factors hypothesized to influence the risk of type 1 diabetes tend to be more common in lower socioeconomic groups, including smoking in pregnancy, obesity, infections, and short duration of breastfeeding [6-8]. We recently reviewed the literature and found few high-quality studies relating socioeconomic status and childhood-onset type 1 diabetes [9]. Many studies used area-level indicators of socioeconomic status at the time of diagnosis, a design susceptible to multiple biases [10]. Few previous studies accounted for confounding by ethnicity or immigration background.

Furthermore, while parental occupation has been investigated in relation to childhood onset-type 1 diabetes as an indicator of social status, some occupations involve exposures that may be related to the aetiology of type 1 diabetes. However, our comprehensive review did not identify any large-scale studies with individual level data on maternal or paternal occupation in relation to risk of type 1 diabetes [9]. In a setting where little is known about potential environmental risk factors, we believe investigating potential differences in risk of type 1 diabetes by parental occupations may be informative regarding potential environmental exposures. Examples from other fields include asthma and allergies in children of farmers, childhood cancer risk by parental occupational exposures, and exposure to infections in parents working in schools or health care [11-14].

Nationwide, mandatory registers available in Norway provide a unique opportunity for follow-up for studies of social gradients in incidence of type 1 diabetes in an unselected population. We aimed to examine the potential association of maternal and paternal education and occupation on the incidence of childhood-onset type 1 diabetes in Norway using these linked registers. In addition to assessing socio-economic status, we explored potential association of specific parental occupations.

## Methods

### Study design and population

This was a cohort study using nationwide registers linked at the individual level by the unique personal identification number assigned to all residents in Norway. Participants were children born in Norway from 1974 to 2013 followed for type 1 diabetes diagnosed before 15 years of age. We restricted the analyses to children, and parents born in Norway, to ensure completeness and uniformity of data on parental education and occupation (Figure 1). Two sets of linked individual-level data were included, covering two calendar periods with incident cases of type 1 diabetes diagnosed before age 15 years: 1989-2003 and 2005-2013 (Figure 1, details in Supplementary Information). Children born in Norway born during 1974-2013 were followed for type 1 diabetes, and the education and occupation of their parents were obtained from Statistics Norway.

**Figure 1.**
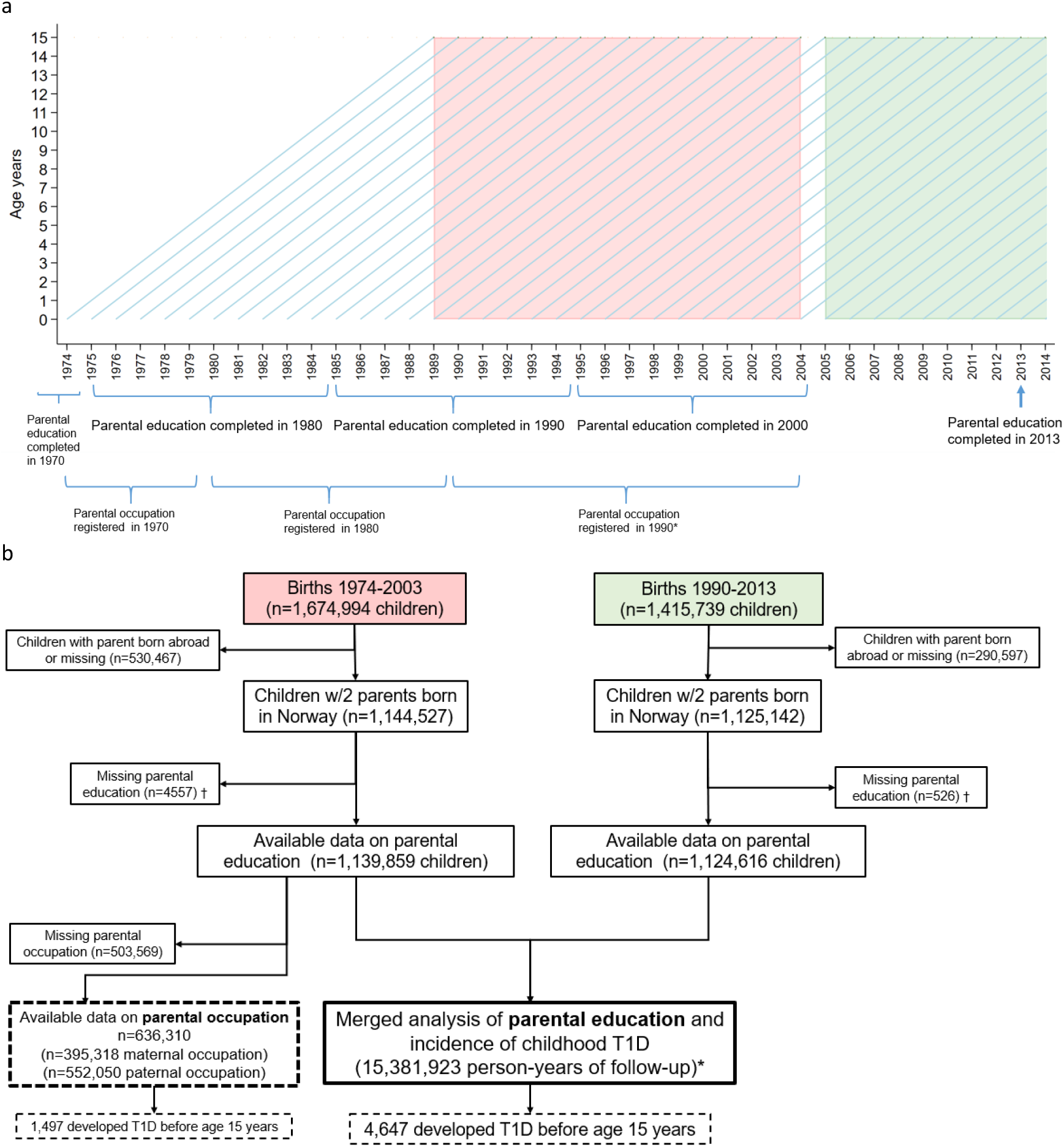
Illustration of study design and formation of analysis sample. a) Birth cohorts, age and timing of parental education and education, and type 1 diabetes (T1D) in children. Incident T1D before age 15 years was ascertained by the Norwegian Childhood Diabetes Registry during 1989-2003 (red box) and by incident use of insulin at least twice according to the Norwegian Prescription Database during 2005-2014 (green box). b) Formation of the analysis sample. * Two sets of linked data were split in one-year intervals of attained age and calendar year. Person-time and incident cases in each age- and calendar year combination were merged to obtain independent contributions from each of the two linked data sets in the merged data for analysis. Detailed explanation in Supplemental Material, p.2. † Missing parental education (both parents).

### Outcome: Type 1 diabetes

Newly diagnosed, clinical type 1 diabetes was the outcome (Figure 1, details in Supplementary Information).

### Parental education

Information on parental education in nine categories was obtained from Statistics Norway, and we used seven categories for the current analyses (see details in Supplementary Information). We used parental education and occupation information collected as close as possible to the time of birth of the child for period 1. For period 2, maternal and paternal education was only available for 2013, as illustrated in Figure 1a. In 1990, information was only available in a representative sample of the population (of parents). We assessed the effect of analysing information from different time-points in robustness analyses. There has been an increase in the education level in the population over time, and the maternal and paternal education were correlated with a Spearman coefficient of 0.43 (Supplementary Information Figure S1).

### Parental occupation

Information on parental occupation was available only in the register linkage for the first study period (Figure 1a). The primary analysis of occupation used 11 categories predefined by Statistics Norway to roughly correspond to socio-economic status. In addition, in an exploratory analysis to assess potential associations of parental occupations with risk of childhood type 1 diabetes, we analysed occupations (two-digit resolution in the Nordic Classification of Occupations) with at least 20 observed cases of type 1 diabetes. Examples of occupations of specific interest were those typically involving social interaction with many children and with infected people. This includes health care workers and teachers. In addition, we were interested in exposure to farm environment.

### Covariates

We included the following covariates in adjusted regression models: Maternal and paternal age at delivery, parity, caesarean section, sex, county of residence at birth of the child obtained from the Medical Birth Registry of Norway (age and calendar period was also included as described below). Maternal smoking during pregnancy was available from the Medical Birth Registry of Norway for births from 1999 (Period 2) and additionally adjusted for in robustness analyses. Maternal type 1 diabetes was available with less than complete coverage for mothers in period 1 and from the Medical Birth Registry for births from 1999 onwards (Period 2). Adjustment for maternal type 1 diabetes was also done in robustness analyses. A conceptual model for how different variables may be linked are shown in Supplementary Information Figure S2.

### Statistical analysis

We split individual follow-up times into 1-year categories of attained age and calendar year stratified by exposure and covariates. We estimated adjusted incidence rate ratios (aIRR) with 95% confidence intervals using Poisson regression. We modelled attained age and calendar period flexibly with restricted cubic splines [15, 16]. For maternal and paternal education, we merged the cross-classified person-time and outcome counts from the two linked datasets. Data were analysed as complete case. The primary statistical test was a test for trend across categories, and we evaluated non-linear associations by categorical analysis and by evaluating a square term for exposure. Analyses were done using STATA version 16 (StataCorp, College Station, TX).

### Ethics

The study was approved by Regional Committee for Medical and Health Research Ethics, South-East Norway C (reference 2012/3 data set for study period 1) and Regional Committee for Medical and Health Research Ethics, South-East Norway B (reference 2010/2583, data set for study period 2).

## Results

During 15,381,923 person-years of follow-up, 4647 children developed type 1 diabetes (Figure 1). The incidence rate was higher in the most recent period, and slightly higher in boys than in girls (Table 1, as previously reported [17]).

**Table 1.** Characteristics of cohort giving rise to incident cases of type 1 diabetes during two periods. Children born in Norway to Norwegian-born parents.

|  |  | Period 1989-2003 |  |  | Period 2005-2013 |  |  |
| --- | --- | --- | --- | --- | --- | --- | --- |
|  |  | Person-years † | T1D cases | Incidence rate* | Person-years† | T1D cases | Incidence rate* |
|  | Total | 9192165 | 2325 | 25.3 (24.3 - 26.3) | 6292314 | 2338 | 37.2 (35.7 - 38.7) |
| Child's sex | Boys | 4723688 | 1242 | 26.3 (24.9 - 27.8) | 3228802 | 1260 | 39.0 (36.9 - 41.2) |
|  | Girls | 4468300 | 1083 | 24.2 (22.8 - 25.7) | 3063513 | 1078 | 35.2 (33.1 - 37.4) |
| Maternal age at delivery | <20 | 269577 | 79 | 29.3 (23.5 - 36.5) | 163294 | 61 | 37.4 (29.1 - 48.0) |
|  | 20-24 | 2261783 | 544 | 24.1 (22.1 - 26.2) | 1013357 | 388 | 38.3 (34.7 - 42.3) |
|  | 25-29 | 3661699 | 915 | 25.0 (23.4 - 26.7) | 2187115 | 835 | 38.2 (35.7 - 40.9) |
|  | 30-34 | 2272315 | 589 | 25.9 (23.9 - 28.1) | 1997983 | 723 | 36.2 (33.6 - 38.9) |
|  | ≥ 35 | 726791 | 198 | 27.2 (23.7 - 31.3) | 930538 | 331 | 35.6 (31.9 - 39.6) |
| Parity (birth order) | 1st | 3889582 | 990 | 25.5 (23.9 - 27.1) | 2568947 | 909 | 35.4 (33.2 - 37.8) |
|  | 2nd | 3419412 | 870 | 25.4 (23.8 - 27.2) | 2282775 | 861 | 37.7 (35.3 - 40.3) |
|  | 3rd | 1456259 | 348 | 23.9 (21.5 - 26.5) | 1063028 | 421 | 39.6 (36.0 - 43.6) |
|  | 4th | 329224 | 93 | 28.2 (23.1 - 34.6) | 276206 | 115 | 41.6 (34.7 - 50.0) |
|  | ≥5th | 97688 | 24 | 24.6 (16.5 - 36.7) | 101358 | 32 | 31.6 (22.3 - 44.6) |
| Caesarean section | No | 8146319 | 2041 | 25.1 (24.0 - 26.2) | 5379958 | 1974 | 36.7 (35.1 - 38.3) |
|  | Yes | 1045846 | 284 | 27.2 (24.2 - 30.5) | 912357 | 364 | 39.9 (36.0 - 44.2) |
| Paternal age at delivery | <25 | 1192733 | 315 | 26.4 (23.6 - 29.5) | 568355 | 214 | 37.7 (32.9 - 43.1) |
|  | 25-29 | 3288251 | 810 | 24.6 (23.0 - 26.4) | 1690550 | 661 | 39.1 (36.2 - 42.2) |
|  | 30-34 | 3046489 | 780 | 25.6 (23.9 - 27.5) | 2170022 | 780 | 35.9 (33.5 - 38.6) |
|  | 35-39 | 1313297 | 348 | 26.5 (23.9 - 29.4) | 1253151 | 481 | 38.4 (35.1 - 42.0) |
|  | ≥40 | 351394 | 72 | 20.5 (16.3 - 25.8) | 573854 | 193 | 33.6 (29.2 - 38.7) |

\* Rate per 100,000 person-years (95% confidence interval). T1D: Type 1 diabetes. T1D cases: Number of incident cases during follow-up.
† Total cohort size was 1,139,859 children in period 1989-2003 and 1,125,142 children in period 1990-2013. Many children contributed with person-years in both periods (See also Figure 1a).

### Maternal or paternal education in relation to risk of type 1 diabetes

Maternal education showed an overall significant inverse association with incidence of type 1 diabetes (p[trend]= 0.043), which was non-linear (p[squared term]=0.027; six degree of freedom likelihood ratio test: p=0.011). There was no difference between level of maternal education among the four lower categories, but a decreasing incidence from the fourth to seventh category (Figure 2). For instance, those with mothers with master’s degree had an 18% lower incidence compared to those who only completed upper secondary education (aIRR=0.82 95% confidence interval: 0.70 - 0.95) (Figure 2, further details Supplementary Information Table S1). These results were consistent in period 1 (1989-2004) and 2 (2005-2013) (Supplementary Information Table S2). In robustness analyses, results remained essentially unchanged or even stronger after adjusting for maternal smoking in the sub-cohort with available data (Supplementary Information Figure S3). Characteristics of those with available maternal smoking data were largely similar to that in the whole cohort (Supplementary Information Table S3). Results were also similar after adjusting for maternal type 1 diabetes (Supplementary Information Table S4).

**Figure 2.**
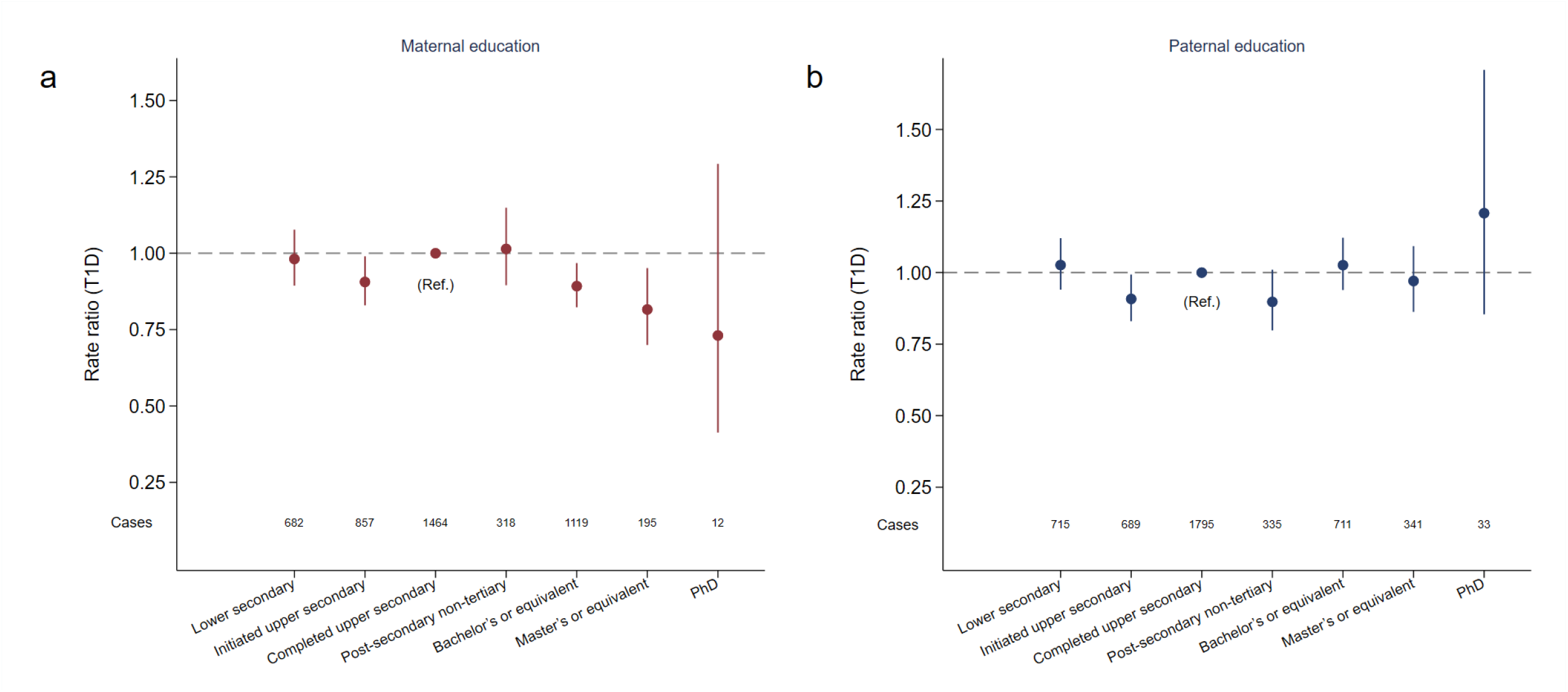
Parental education and incidence of type 1 diabetes (T1D) a) Maternal education and b) Paternal education. Nationwide cohort of children in Norway of whom 4647 developed T1D before 15 years of age during 1989-2013. Incidence rate ratios (aIRR) were adjusted for parental age at delivery, parity, caesarean section, child’s sex, county of residence, age and calendar period. Vertical lines represent 95% confidence intervals for the aIRRs. Statistical test for trend: p=0.043 for maternal education and p=0.83 for paternal education. There was significant deviation from linearity for maternal education (square term p=0.027; six degree of freedom likelihood ratio test for maternal education: p=0.011. Details regarding numbers in each category, absolute incidence rates and characteristics for each category of maternal education are shown in Supplemental Table S1.

Paternal education was not significantly associated with incidence of type 1 diabetes overall (Figure 2) or in any of the two periods (Supplementary Information Table S2). Robustness analyses showed that associations of maternal or paternal education with type 1 diabetes was similar if we used education at different time points (Supplementary Information Table S5).

### Parental occupation and incidence of type 1 diabetes

There was a suggestive non-linear association between maternal occupation categorized according to socioeconomic status, and risk of type 1 diabetes (Figure 3a). Children of mothers with elementary occupations (for example: hotel, restaurant, household worker, cleaners) had a 23% lower risk of type 1 diabetes compared to children with mothers in professional occupations (aIRR=0.77, 95% CI: 0.63-0.94). Paternal occupation was not significantly associated with type 1 diabetes (Figure 3b). Additional adjustment for parental education gave similar results (Supplementary Information Figure S4). Maternal and paternal occupation was available only for a subset in some of the early censuses, and characteristics were similar for those with and without available data on maternal and paternal occupation (Supplementary Information Table S6).

**Figure 3.**
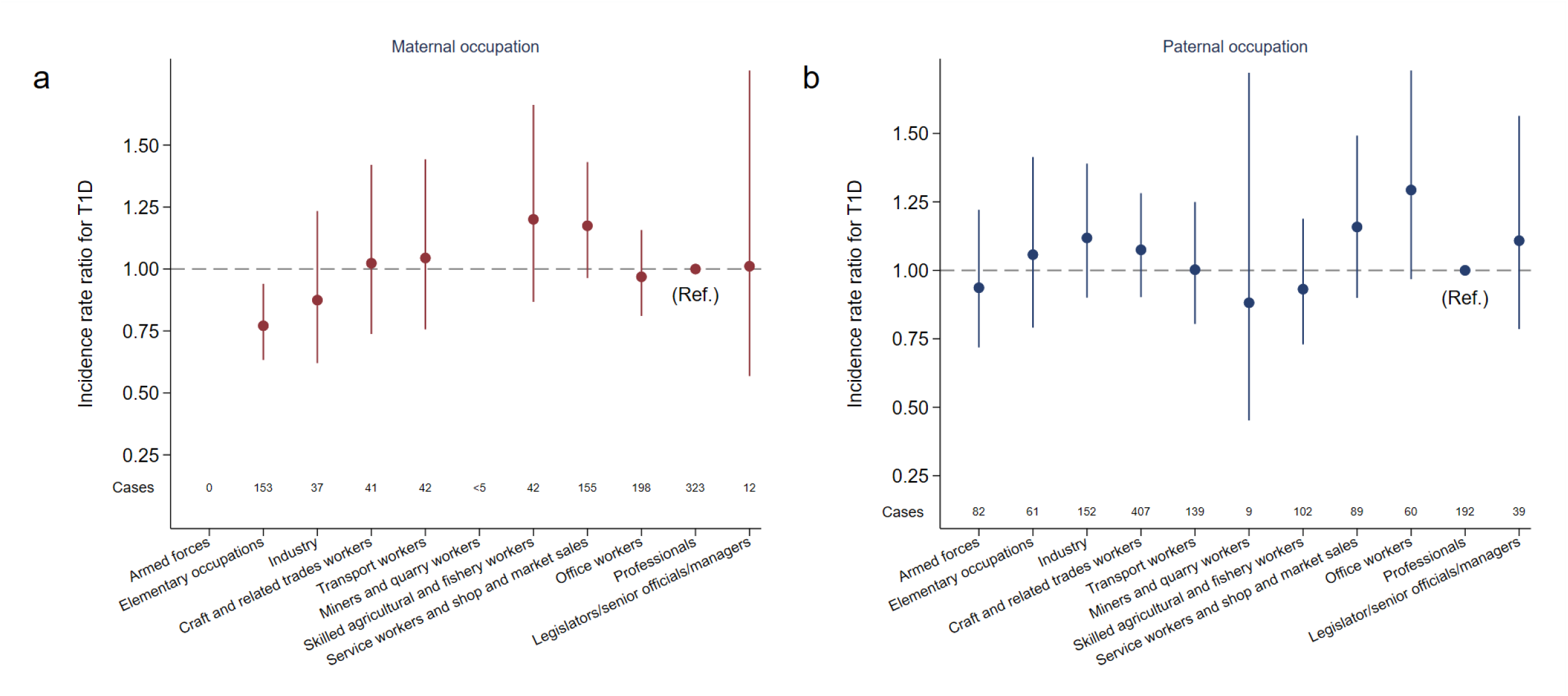
Parental occupation and incidence of type 1 diabetes (T1D) a) Maternal occupation, and b) paternal occupation. Nationwide cohort of children in Norway of whom 1,497 developed T1D before 15 years of age during 1989-2003. Incidence rate ratios (aIRR) were adjusted for parental age at delivery, parity, caesarean section, child’s sex, county of residence, age and calendar period. Vertical lines represent 95% confidence intervals (CI) for the aIRRs. The aIRR for mothers who worked in mining and quarry industry was not shown in the figure due to low number of cases, but it was 5.65, 95% CI: 1.41-22.7. Likelihood ratio test for overall association (10 degree of freedom): p=0.017 for maternal occupation and p=0.64 for paternal occupation. Additional adjustment for maternal education gave similar results (Figure S4).

Finally, we explored the incidence of type 1 diabetes across more detailed occupational categories including some of specific interest: Children of mothers working as nurses or teachers are typically exposed to social interaction including people with infectious diseases, and farmers. However, both teachers and farmers ranked average among the common maternal occupations (Supplementary Information Table S7). The subcategories that were elementary also tended to show lower risk (occupation code 91, 92 and 93 in Supplementary Information Table S7). Hotel, restaurant, and related workers (code 91) ranked lowest for maternal occupations. Subcategories of paternal occupations showed no consistent pattern.

## Discussion

We found that higher maternal education was associated with a slightly lower risk of childhood-onset type 1 diabetes risk, while paternal education was not. Maternal elementary occupations were associated with lower risk of type 1 diabetes, but paternal occupations did not show any consistent association with type 1 diabetes.

Major strengths of our study include the mandatory nationwide register-based design with individual-level information, and large sample size. We accounted for parental country of birth [18], maternal smoking in pregnancy [19], and maternal age, potentially important confounders not adjusted for in many previous studies.

Our recent literature review identified methodological weaknesses in the majority of previous studies in this field, particularly semi-ecologic and case-control studies [9]. Two previous studies with good methodology showed signs of non-linear associations between parental education and risk of childhood type 1 diabetes [20, 21]. These prompted us to analyse socioeconomic status variables in multiple categories to assess “dose-response” relationships. The two previous studies, from Turin and USA, respectively, found lowest risk associated with low education [20, 21]. In our study, we found lowest risk with high maternal education, and statistical evidence for an inverse U-shaped relationship between maternal socioeconomic status and type 1 diabetes. Two other large Scandinavian cohort studies also found inverse U-shaped associations consistent with our result [22, 23]. The lack of association between paternal education and childhood type 1 diabetes in our study was confirmed in the only previous cohort study with such data [22].

Our study is to our knowledge the first large scale study to systematically investigate parental occupations in relation to risk of type 1 diabetes. A previous case-control study reported that parental occupations involving exposure to children was associated with lower risk of type 1 diabetes, although specific occupations were not shown [13]. We chose to study specific parental occupations such as healthcare workers and teachers but found no association with childhood-onset type 1 diabetes in our cohort. Growing up on a farm has been associated with different gut microbiotas and lower risk of asthma and some allergies [24]. Three previous studies investigated farming environment in relation to risk of type 1 diabetes. None of these reported significant associations, but sample size and study design render these studies inconclusive [25-27]. Having parents working on farms was not associated with incidence of type 1 diabetes in our study.

Indicators of socioeconomic status are unlikely to be directly causally related to type 1 diabetes. Interpretation is complex and likely context dependent. However, we believe our study contribute with important information in a setting where few environmental factors are established as risk factors and several exposures hypothesized to be related to type 1 diabetes have shown to be associated with socioeconomic status. Together with the few previous studies with good methodological quality, our results show that socioeconomic status does not have a very strong relationship with type 1 diabetes. If anything, maternal indicators of socioeconomic status seem more likely relevant than do paternal indicators. the underlying suggestive non-linear associations we report may partially explain the inconsistencies among previous studies. This is because many previous studies had used only two or three categories of socioeconomic status and different (often unreported) cut-offs between categories.

Our findings also have practical implications for future studies. First, given the relatively weak and inconsistent associations between socioeconomic status and type 1 diabetes, adjustment for socioeconomic status when studying other exposures may not be essential in many settings. Second, we recommend including clearly defined individual-level socioeconomic variables separately for the mother and the father. One of the previous studies with otherwise good methodology used the highest of the father’s or mother’s education, which complicates interpretation [20]. We speculate that the association with maternal and not paternal education may be driven by intrauterine or early postnatal environmental exposures typically influenced by maternal lifestyle and decisions, such as infant and childhood nutrition and hygiene.

Our study has some limitations. We did not have information on family income. However, our recent review found few other studies and no consistent association between family income and risk of type 1 diabetes. While we focused our analysis on parental education and occupation at the time of the child’s birth to avoid potential reverse causality, we did not always have socioeconomic status at the exact year of birth of the child, and data on socioeconomic status and confounders were missing for some periods. On the other hand, our robustness analyses suggested that this did was unlikely to materially influence our results. We cannot generalize our findings to immigrants because information on education and occupation in the registries is incomplete for immigrants. On the other hand, the proportion of immigrants in Norway is too small to have allowed robust analyses in this subgroup even if we had valid information on socioeconomic status. While our large sample size allowed for analysis of parental education and occupation in multiple categories, we decided a priori not to stratify our analyses by sub-groups, because such analyses would have low statistical power and complicate interpretation [28]. Finally, it may be argued that Nordic countries have a relatively egalitarian society with universal and free access to health care and education, and that limited social inequality in health is to be expected. However, clear social gradients in adult mortality and self-rated health have been found in the Nordic countries [29]. Furthermore, higher maternal education has been robustly associated with lower childhood adiposity [8] and lower childhood mortality in the Nordic countries [30]. We therefore believe that differences in socioeconomic status within Norway are not negligible and that we should be able to detect associations with our large sample size, if there truly is a relationship.

## Conclusions

In conclusion, our results suggested associations between maternal socioeconomic status, with a slightly lower risk of type 1 diabetes in children of mothers with higher education. Paternal socioeconomic status was not associated with type 1 diabetes in children.

## Availability of data

Norwegian data protection legislation and Act on medical and health research do not allow individual level patient data to be shared by the authors. However, all data are accessible to authorised researchers after ethical approval and application to the registries via https://helsedata.no/ administered by the Directorate of eHealth, and the Norwegian Childhood Diabetes Registry.

## Author Contributions

LCS and GJ conceptualized the study and research question. GJ, LCS and SEH were responsible for data acquisition, planning, and funding. LCS and PLDR conceptualized the detailed study design. PLDR, GT and IJB carried out the data preparation. PLDR and LCS did the statistical analyses with input and quality control by GT. All authors contributed to the interpretation of results. PLDR wrote the first draft of the manuscript and subsequent revisions. All authors critically revised the paper for important intellectual content and approved the final version. PLDR and LCS had full access to all the data in the study and take responsibility for the integrity of the data and the accuracy of the data analysis.

