## Supplementary Information for "Parental education and occupation in relation to risk of childhood type 1 diabetes: nationwide cohort study"

### Supplementary Methods

#### The Norwegian education and occupation system and data collection

Upper secondary education is compulsory in Norway, and the duration of compulsory education has varied from seven years up to 1969, nine years during 1969-1997, and 10 years after 1997. Number of years of education for a given level may therefore vary by calendar year. The Norwegian standard for educational groups (NUS) started in 1970, with different versions in 1973 and 1989, and data on the population was collected by Statistics Norway in decennial censuses in 1970, 1980, and 1990 (representative subsample in 1990). Parental educational data closest to the birth for the child was used in the analysis for period 1 (1989-2003), while data from 2013 was used for period 2 (2005-2013) as explained the main text methods section and main Figure 1b. The educational codes are explained below:

| Original statistics Norway category | Statistics Norway coding | Years of education* | Categories used in the statistical analysis with description |
|---|---|---|---|
| | 0 | - | 1.Lower secondary |
| Compulsory education | 1 | 1-7 years | |
| | 2 | 8-10 years | |
| Middle education | 3 | Over 8 years | 2. Initiated upper secondary (but not completed) |
| | 4 | 10-13 years | 3. Completed upper secondary (high school or vocational education) |
| | 5 | 13-15 years | 4. Post-secondary non-tertiary (started additional education after upper secondary, but not completed any higher degree) |
| High education | 6 | 15-17 years | 5. Bachelor's degree or equivalent |
| | 7 | 17-19 years | 6. Master's degree or equivalent |
| | 8 | ≥ 20 years | 7. PhD level |
| | 9 | Not provided / missing | Not included in the analyses |

* Number of years has varied over time as explained in the text. In early years those who did not pass and exam after 7 years of compulsory education were coded with zero. For most of the period covered, all residents received level 1 compulsory education (6-7 years) with no requirement to pass a test.

Nordic Classification of Occupations for parents were similarly available for these time points in period 1 (1989-2003) https://www.nb.no/nbsok/nb/8d2a147fbb144dbb1fa40670315cb966?lang=no#0

#### Operational definition of newly diagnosed type 1 diabetes

The study outcome, newly diagnosed clinical type 1 diabetes before age 15 years, was defined using registries, with slightly different operational definitions for the two study periods. For 1989-2003 (the first linked data set), used the Norwegian Childhood Diabetes Registry, a nation-wide medical quality registry with nearly complete coverage which participates in EURODIAB incidence studies (1). This is run by paediatricians specializing in diabetes care throughout Norway. Date of diagnosis defined as the day of the first insulin injection (2). For the period 2005-2013, type 1 diabetes was defined at the first date of dispensing insulin prescriptions according to the Norwegian prescription database (Anatomical Therapeutic Chemical Classification code A10A). We required at least two separate dispensed insulin prescriptions, and no use of sulfonylureas to avoid other types of diabetes. For individuals starting with insulin in 2013, we allowed for the second dose to be dispensed in 2014. To exclude prevalent cases of type 1 diabetes in the second period, we required no use of insulin in 2004, and we were thus not able to include incident type 1 diabetes cases in in 2004 (see also Main Figure 1).

#### Methods for combining two data sets for maternal and paternal education

We included two datasets, each containing individual linkage of nationwide registries with all individuals who were diagnosed with type 1 diabetes before age 15 years in the Norwegian population during 1989-2003 (pink section in figure), and 2005-2013 (green section in figure) inclusive. These datasets had previously been used for different purposes, but the study of social inequality in risk of type 1 diabetes was an aim of both projects (3, 4).

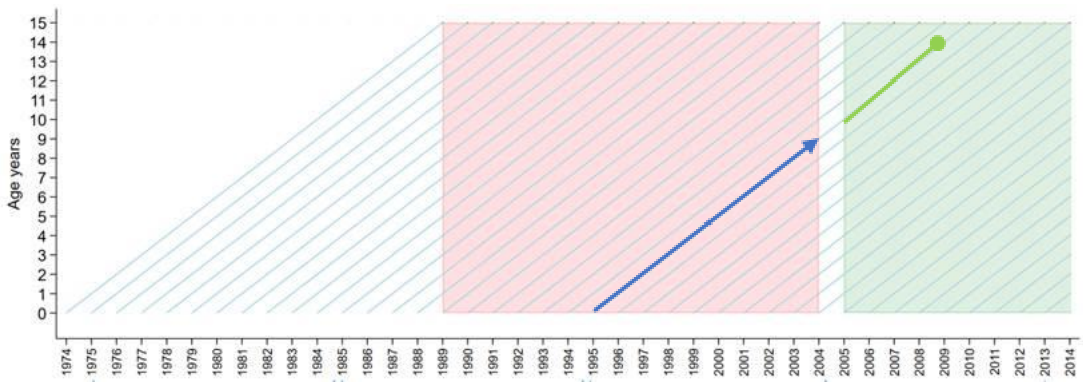

The figure (Lexis diagram) shows that children diagnosed in each period could be born before the period of type 1 diabetes ascertainment (a child diagnosed in 1989 could be born 1974-1989, and a child diagnosed in 2005 could be born 1990-2005). In individual-level cohort analysis of each of the two datasets, person-time at follow-up for children born before the periods of ascertainment was not counted before the child was “at risk” of being identified with newly diagnosed type 1 diabetes in the respective pink and green periods (this is often called left truncated survival data, see also additional explanation for the white segment in 2004 below). Children born 1990-2003 appeared in both datasets and contribute person-time at risk of incident type 1 diabetes in different calendar periods at different ages.

How did we ensure that a child was not counted twice? For example, a girl born 1st of January 1995 contributed nine (person-) years of follow-up (1995 – 2004) in the first period (blue thick arrow across the pink area), until her 9th birthday, and she did not develop type 1 diabetes during this first period. From 2005 to 2008, she contributed another four person-years of follow-up, from age 11 until the 14th birthday when she was diagnosed with type 1 diabetes (indicated by the circle at the end of the thick green line). Because both datasets included information about date of birth, date of diagnosis of type 1 diabetes, exposures and covariates, we could cross-classify each individual child’s person-time under follow-up simultaneously by the two time-dependent variables age and calendar period (using the “stsplit” function in Stata we split the person-time for each individual in the two separate datasets in 1-year categories of age and calendar period and category of exposure and covariate). Person-time and number of incident cases were further stratified by exposure and covariates using the “stptime” function in Stata. This allowed us to add the person-time of follow-up (and number of incident cases of type 1 diabetes) for all children in each stratum of age, period, exposure, and covariate from the two datasets, because any child that appeared in both data sets appeared in a different stratum of age- and calendar period in the two data sets. Thus, we could analyse the resulting table of cross-classified person-time and cases of incident type 1 diabetes using Poisson regression just as if we had a single dataset covering the entire period with type 1 diabetes diagnosed from 1989-2013 (5). We modelled these data across the two periods with Poisson regression as described in the main methods section with restricted cubic splines for age and period, which produces results equivalent to that of Cox regression (6). Finally, we could not include follow-up time of incident cases for 2004 (the white vertical segment in the Lexis diagram). This is because newly diagnosed type 1 diabetes in the second period was done using the Norwegian Prescription database and individual using insulin in 2004 contained both incident and prevalent cases of type 1 diabetes. To ensure we had incident (newly diagnosed) cases, we required that individuals had not dispensed insulin prescriptions through 2004.

**Supplementary methods references**

1. Patterson CC, Gyurus E, Rosenbauer J, Cinek O, Neu A, Schober E, et al. Trends in childhood type 1 diabetes incidence in Europe during 1989-2008: evidence of non-uniformity over time in rates of increase. Diabetologia. 2012;55(8):2142-7.
2. Joner G, Stene LC, Søvik O. Nationwide, prospective registration of type 1 diabetes in children aged< 15 years in Norway 1989–1998: no increase but significant regional variation in incidence. Diabetes Care. 2004;27(7):1618-22.
3. Ruiz PLD, Tapia G, Bakken IJ, Haberg SE, Hungnes O, Gulseth HL, et al. Pandemic influenza and subsequent risk of type 1 diabetes: a nationwide cohort study. Diabetologia. 2018;61(9):1996-2004.
4. Stene LC, Magnus P, Lie RT, Søvik O, Joner G, Norwegian childhood Diabetes Study Group. Birth weight and childhood onset type 1 diabetes: population based cohort study. BMJ. 2001;322(7291):889-92.
5. Carstensen B. Age-period-cohort models for the Lexis diagram. Stat Med. 2007;26(15):3018-45.
6. Royston P, Lambert PC. Flexible parametric survival analysis using stata: beyond the Cox model. College Station, Texas: Stata Press; 2011.

### Supplementary results: Tables

#### Table S1. Number of incident cases of type 1 diabetes, absolute incidence rates and characteristics for each category of maternal education*

#### Period 1989-2003

| | | Lower secondary or less (up to 10 years) | Initiated upper secondary (over 11 years) | Upper secondary completed (13 years) | Post-secondary non-tertiary (14 years) | Bachelor's or equivalent level (17 years education) | Master's or equivalent level (19 years education) | PhD (20 years) |
|---|---|---|---|---|---|---|---|---|
| | Total | 313, 27.9 (25.0 - 31.2) | 707, 25.0 (23.2 - 26.9) | 723, 25.9 (24.1 - 27.9) | 253, 29.0 (25.6 - 32.8) | 272, 21.5 (19.1 - 24.2) | 43, 19.5 (14.5 - 26.3) | <5, 17.2 (2.4 - 122.1) |
| **Child's sex** | Boys | 172, 29.8 (25.7 - 34.6) | 374, 25.8 (23.3 - 28.5) | 386, 26.9 (24.4 - 29.7) | 133, 29.6 (24.9 - 35.0) | 145, 22.4 (19.0 - 26.3) | 24, 21.0 (14.1 - 31.3) | <5, 34.1 (4.8 - 241.8) |
| | Girls | 141, 25.9 (21.9 - 30.5) | 333, 24.1 (21.7 - 26.9) | 337, 24.9 (22.4 - 27.7) | 120, 28.4 (23.7 - 33.9) | 127, 20.6 (17.3 - 24.5) | 19, 17.9 (11.4 - 28.1) | - |
| **Maternal age at delivery (years)** | <20 | 25, 25.2 (17.0 - 37.2) | 30, 31.9 (22.3 - 45.6) | 13, 36.5 (21.2 - 62.8) | - | - | - | - |
| | 20-24 | 113, 26.3 (21.9 - 31.6) | 166, 19.4 (16.7 - 22.6) | 221, 27.1 (23.8 - 31.0) | 29, 38.6 (26.8 - 55.5) | 13, 18.7 (10.8 - 32.1) | 12, 18.4 (10.5 - 32.4) | - |
| | 25-29 | 100, 29.1 (23.9 - 35.4) | 270, 25.5 (22.6 - 28.7) | 324, 25.8 (23.1 - 28.7) | 94, 24.6 (20.1 - 30.1) | 115, 21.3 (17.7 - 25.6) | 25, 22.8 (15.4 - 33.7) | <5, 34.9 (4.9 - 247.6) |
| | 30-34 | 58, 32.0 (24.7 - 41.4) | 187, 30.4 (26.3 - 35.1) | 132, 23.7 (20.0 - 28.1) | 85, 27.3 (22.1 - 33.7) | 101, 20.9 (17.2 - 25.3) | 6, 14.0 (6.3 - 31.1) | - |
| | ≥ 35 | 17, 25.2 (15.7 - 40.5) | 54, 25.8 (19.7 - 33.6) | 33, 26.3 (18.7 - 37.0) | 45, 43.7 (32.7 - 58.6) | 43, 25.1 (18.6 - 33.8) | 19, 19.4 (12.4 - 30.5) | - |
| **Parity** | 1st | 126, 29.9 (25.1 - 35.6) | 254, 24.5 (21.7 - 27.7) | 355, 26.1 (23.5 - 28.9) | 114, 30.5 (25.4 - 36.6) | 109, 20.0 (16.6 - 24.1) | 17, 20.4 (12.7 - 32.9) | <5, 44.3 (6.2 - 314.6) |
| | 2nd | 111, 27.8 (23.0 - 33.4) | 267, 24.3 (21.5 - 27.4) | 282, 27.8 (24.8 - 31.3) | 83, 25.2 (20.3 - 31.3) | 109, 23.0 (19.1 - 27.8) | 6, 18.6 (8.4 - 41.4) | - |
| | 3rd | 54, 25.6 (19.6 - 33.4) | 135, 25.6 (21.6 - 30.3) | 67, 19.7 (15.5 - 25.0) | 43, 31.7 (23.5 - 42.7) | 43, 21.5 (16.0 - 29.0) | - | - |
| | 4th | 16, 25.0 (15.3 - 40.8) | 44, 34.5 (25.6 - 46.3) | 18, 29.4 (18.5 - 46.7) | 7, 25.1 (12.0 - 52.7) | 8, 20.4 (10.2 - 40.8) | <5, 85.8 (12.1 - 609.1) | - |
| | ≥5th | 6, 23.3 (10.5 - 51.8) | 7, 17.3 (8.3 - 36.4) | <5, 7.0 (1.0 - 49.8) | 6, 91.2 (41.0 - 202.9) | <5, 35.7 (11.5 - 110.7) | 38, 19.8 (14.4 - 27.2) | <5, 20.0 (2.8 - 141.8) |
| **Caesarean section** | No | 263, 26.3 (23.3 - 29.7) | 624, 24.9 (23.0 - 26.9) | 645, 26.0 (24.1 - 28.1) | 216, 27.9 (24.4 - 31.8) | 243, 21.8 (19.2 - 24.7) | 5, 17.4 (7.2 - 41.8) | - |
| | Yes | 50, 40.8 (30.9 - 53.8) | 83, 25.7 (20.8 - 31.9) | 78, 25.0 (20.0 - 31.2) | 37, 37.8 (27.4 - 52.2) | 29, 19.2 (13.3 - 27.6) | - | - |
| **Paternal age at) delivery (years** | <25 | 72, 26.2 (20.8 - 33.0) | 104, 23.2 (19.1 - 28.1) | 112, 30.9 (25.7 - 37.2) | 12, 35.9 (20.4 - 63.2) | 7, 20.4 (9.7 - 42.7) | 7, 16.9 (8.0 - 35.4) | - |
| | 25-29 | 110, 26.8 (22.2 - 32.3) | 240, 22.8 (20.1 - 25.9) | 291, 25.8 (23.0 - 28.9) | 82, 29.6 (23.8 - 36.7) | 77, 21.7 (17.4 - 27.1) | 25, 24.8 (16.8 - 36.7) | <5, 47.1 (6.6 - 334.0) |
| | 30-34 | 88, 30.9 (25.1 - 38.1) | 246, 28.4 (25.1 - 32.2) | 217, 24.3 (21.3 - 27.7) | 96, 26.5 (21.7 - 32.4) | 105, 20.1 (16.6 - 24.3) | 10, 17.1 (9.2 - 31.8) | - |
| | 35-39 | 33, 27.6 (19.6 - 38.8) | 97, 26.5 (21.8 - 32.4) | 91, 27.8 (22.7 - 34.2) | 50, 30.1 (22.8 - 39.7) | 67, 25.0 (19.7 - 31.8) | <5, 5.5 (0.8 - 39.1) | - |
| | ≥40 | 10, 31.0 (16.7 - 57.6) | 20, 20.3 (13.1 - 31.5) | 12, 15.4 (8.7 - 27.1) | 13, 37.9 (22.0 - 65.4) | 16, 18.3 (11.2 - 29.9) | 43, 19.5 (14.5 - 26.3) | <5, 17.2 (2.4 - 122.1) |

**Table S1, continued. Period 2005-2013**

| | | **Started lower secondary (10 years education)** | **Upper secondary (over 11 years)** | **Upper secondary completed (13 years)** | **Post-secondary non-tertiary (14 years)** | **Bachelor's or equivalent level (17 years education)** | **Master's or equivalent level (19 years education)** | **PhD (20 years)** |
|---|---|---|---|---|---|---|---|---|
| | Total | 369, 38.9 (35.1 - 43.1) | 150, 41.4 (35.3 - 48.6) | 741, 40.6 (37.8 - 43.6) | 65, 40.0 (31.4 - 51.0) | 847, 34.9 (32.6 - 37.3) | 152, 29.5 (25.2 - 34.6) | 11, 27.5 (15.2 - 49.6) |
| **Child's sex** | Boys | 213, 43.6 (38.1 - 49.9) | 80, 42.9 (34.5 - 53.5) | 412, 44.0 (39.9 - 48.4) | 41, 49.2 (36.2 - 66.8) | 433, 34.8 (31.7 - 38.2) | 871, 26.9 (21.3 - 34.0) | 8, 38.5 (19.3 - 77.1) |
| | Girls | 156, 33.9 (29.0 - 39.6) | 70, 39.8 (31.5 - 50.3) | 329, 37.0 (33.2 - 41.2) | 24, 30.3 (20.3 - 45.2) | 414, 35.0 (31.7 - 38.5) | 81, 32.3 (25.9 - 40.1) | <5, 15.6 (5.0 - 48.2) |
| **Maternal age at delivery (years)** | <20 | 27, 33.3 (22.9 - 48.6) | <5, 95.3 (30.7 - 295.4) | 21, 40.6 (26.5 - 62.2) | - | 10, 43.1 (23.2 - 80.1) | - | - |
| | 20-24 | 90, 35.2 (28.6 - 43.2) | 21, 43.3 (28.2 - 66.4) | 161, 39.6 (34.0 - 46.3) | 13, 66.3 (38.5 - 114.2) | 97, 37.2 (30.5 - 45.4) | 5, 25.4 (10.6 - 61.1) | - |
| | 25-29 | 114, 41.3 (34.3 - 49.6) | 51, 43.3 (32.9 - 57.0) | 297, 43.8 (39.1 - 49.1) | 20, 36.4 (23.5 - 56.4) | 302, 33.5 (29.9 - 37.5) | 47, 32.0 (24.0 - 42.6) | <5, 41.7 (15.6 - 111) |
| | 30-34 | 93, 43.3 (35.3 - 53.0) | 42, 36.6 (27.0 - 49.5) | 179, 36.3 (31.4 - 42.0) | 22, 39.1 (25.7 - 59.4) | 316, 36.5 (32.7 - 40.8) | 64, 27.4 (21.4 - 35.0) | 5, 27.7 (11.5 - 66.5) |
| | ≥ 35 | 45, 37.2 (27.8 - 49.9) | 33, 42.2 (30.0 - 59.4) | 83, 42.0 (33.9 - 52.1) | 10, 34.1 (18.4 - 63.4) | 122, 32.2 (27.0 - 38.5) | 36, 31.9 (23.0 - 44.3) | <5, 17.3 (4.3 - 69.2) |
| **Parity** | 1st | 122, 34.8 (29.2 - 41.6) | 42, 39.9 (29.5 - 54.0) | 291, 38.4 (34.3 - 43.1) | 29, 46.3 (32.2 - 66.6) | 345, 33.3 (29.9 - 37.0) | 77, 32.5 (26.0 - 40.7) | <5, 17.3 (5.6 - 53.7) |
| | 2nd | 126, 40.0 (33.6 - 47.6) | 59, 43.2 (33.5 - 55.8) | 292, 43.1 (38.5 - 48.4) | 26, 41.6 (28.4 - 61.2) | 306, 34.4 (30.8 - 38.5) | 46, 24.8 (18.6 - 33.2) | 5, 32.8 (13.6 - 78.8) |
| | 3rd | 81, 44.6 (35.9 - 55.5) | 34, 41.2 (29.4 - 57.6) | 122, 41.3 (34.6 - 49.3) | 7, 24.5 (11.7 - 51.5) | 155, 39.5 (33.8 - 46.3) | 19, 25.4 (16.2 - 39.9) | <5, 32.5 (8.1 - 130.0) |
| | 4th | 28, 41.5 (28.6 - 60.1) | 11, 41.8 (23.1 - 75.4) | 28, 38.8 (26.8 - 56.2) | <5, 29.1 ( 7.3 - 116.4) | 38, 43.9 (32.0 - 60.4) | 7, 47.4 (22.6 - 99.4) | <5, 92.2 (13.0 - 655) |
| | ≥5th | 12, 34.7 (19.7 - 61.1) | <5, 34.5 (13.0 - 92.0) | 8, 32.6 (16.3 - 65.2) | <5, 49.7 ( 7.0 - 352.6) | <5, 12.3 ( 4.0 - 38.2) | <5, 87.9 (28.3 - 272.4) | - |
| **Caesarean section** | No | 310, 38.6 (34.5 - 43.1) | 123, 40.2 (33.7 - 48.0) | 631, 40.4 (37.4 - 43.7) | 57, 41.5 (32.0 - 53.8) | 710, 34.0 (31.6 - 36.6) | 132, 29.8 (25.1 - 35.3) | 9, 26.3 (13.7 - 50.6) |
| | Yes | 59, 40.7 (31.5 - 52.5) | 27, 47.8 (32.8 - 69.7) | 110, 41.4 (34.3 - 49.9) | 8, 31.9 (16.0 - 63.8) | 137, 40.1 (33.9 - 47.4) | 20, 28.1 (18.1 - 43.6) | <5, 34.3 (8.6 - 137.1) |
| **Paternal age at delivery (years)** | <25 | 64, 36.2 (28.4 - 46.3) | 9, 36.6 (19.0 - 70.3) | 89, 41.9 (34.1 - 51.6) | <5, 9.4 ( 1.3 - 66.9) | 49, 37.1 (28.0 - 49.1) | <5, 18.7 (4.7 - 74.9) | - |
| | 25-29 | 116, 43.6 (36.4 - 52.3) | 45, 50.1 (37.4 - 67.1) | 232, 41.7 (36.7 - 47.5) | 20, 49.9 (32.2 - 77.4) | 214, 33.7 (29.5 - 38.5) | 33, 34.7 (24.6 - 48.8) | <5, 15.5 (2.2 - 109.7) |
| | 30-34 | 91, 35.8 (29.1 - 43.9) | 46, 38.5 (28.8 - 51.4) | 234, 39.3 (34.5 - 44.6) | 20, 34.3 (22.1 - 53.2) | 318, 34.9 (31.2 - 38.9) | 64, 30.2 (23.6 - 38.5) | 5, 32.8 (13.7 - 78.9) |
| | 35-39 | 65, 42.2 (33.1 - 53.8) | 35, 43.0 (30.9 - 59.9) | 138, 43.4 (36.8 - 51.3) | 19, 53.1 (33.9 - 83.3) | 182, 35.2 (30.4 - 40.7) | 38, 28.4 (20.7 - 39.1) | <5, 34.1 (12.8 - 90.9) |
| | ≥40 | 26, 30.5 (20.8 - 44.8) | 15, 32.9 (19.8 - 54.6) | 48, 35.5 (26.8 - 47.2) | 5, 29.4 (12.3 - 70.7) | 82, 36.8 (29.7 - 45.7) | 15, 24.7 (14.9 - 41.0) | <5, 16.3 (2.3 - 115.6) |

* Data in each column: the first number is the number of incident type 1 diabetes cases, and after comma the incidence rate per 100,000 person-years followed by (95% confidence interval). Number of individuals <5 could not be specified because of data protection rules.

**Table S2. Maternal and paternal education and incidence of type 1 diabetes separately in period 1 (1989-2003) and period 2 (2005-2013)**

| **Maternal education** | | | |
|---|---|---|---|
| | **Period 1 (1989-2003)** | **Period 2 (2005-2013)** | **Periods combined** |
| | aIRR (95% CI)* | aIRR (95% CI)* | aIRR (95% CI)* |
| Lower secondary | 1.04 (0.91 - 1.20) | 0.94 (0.83 - 1.06) | 0.98 (0.89 - 1.08) |
| Initiated upper secondary | 0.93 (0.83 - 1.03) | 0.89 (0.74 - 1.06) | 0.91 (0.83 - 0.99) |
| Completed upper secondary (high school or vocational education) | 1 (Reference) | 1 (Reference) | 1 (Reference) |
| Post-secondary non-tertiary (started but not completed any higher degree) | 1.03 (0.89 - 1.19) | 0.95 (0.73 - 1.22) | 1.01 (0.90 - 1.15) |
| Bachelor’s degree or equivalent | 0.86 (0.75 - 1.00) | 0.90 (0.81 - 0.99) | 0.89 (0.82 - 0.97) |
| Master’s degree or equivalent level | 0.79 (0.57 - 1.07) | 0.83 (0.70 - 0.99) | 0.82 (0.70 - 0.95) |
| PhD level | 0.74 (0.10 - 5.27) | 0.74 (0.41 - 1.34) | 0.73 (0.41 - 1.29) |
| **Paternal education** | | | |
| | **Period 1 (1989-2003)** | **Period 2 (2005-2013)** | **Periods combined** |
| | aIRR (95% CI)* | aIRR (95% CI)* | aIRR (95% CI)* |
| Lower secondary | 0.86 (0.75 - 0.98) | 1.06 (0.95 - 1.20) | 0.96 (0.88 - 1.05) |
| Initiated upper secondary | 0.97 (0.87 - 1.08) | 0.93 (0.78 - 1.11) | 0.97 (0.89 - 1.06) |
| Completed upper secondary (high school or vocational education) | 1 (Reference) | 1 (Reference) | 1 (Reference) |
| Post-secondary non-tertiary (started but not completed any higher degree) | 0.88 (0.76 - 1.03) | 0.91 (0.76 - 1.10) | 0.90 (0.80 - 1.01) |
| Bachelor’s degree or equivalent | 0.85 (0.72 - 1.00) | 1.04 (0.93 - 1.16) | 0.97 (0.89 - 1.06) |
| Master’s degree or equivalent level | 0.96 (0.80 - 1.15) | 0.93 (0.79 - 1.08) | 0.93 (0.83 - 1.05) |
| PhD level | 1.11 (0.53 - 2.34) | 1.09 (0.74 - 1.62) | 1.08 (0.76 - 1.52) |

***** Incidence rate ratios (aIRR) for type 1 diabetes, adjusted for maternal age, parity, caesarean section, county of residence, and child’s sex. CI: Confidence interval.

**Table S3. Characteristics of all subjects born 1999-2013 and the subgroup with information on maternal smoking**

| | | All births from 1999-2013 | | | Births 1999-2013 with information on maternal smoking* | | |
|---|---|---|---|---|---|---|---|
| | | **Person-years** | **T1D cases** | **Incidence Rate (per 100 000 PYR)** | **Person-years** | **T1D cases** | **Incidence Rate (per 100 000 PYR)** |
| | Total | 4263652 | 1347 | 31.6 (29.9 - 33.3) | 3468807 | 1108 | 31.9 (30.1 - 33.9) |
| Sex | Boys | 2186566 | 715 | 32.7 (30.4 - 35.2) | 1778712 | 593 | 33.3 (30.8 - 36.1) |
| | Girls | 2077086 | 632 | 30.4 (28.1 - 32.9) | 1690095 | 515 | 30.5 (28.0 - 33.2) |
| Mother age | <20 years | 100210 | 31 | 30.9 (21.8 - 44.0) | 85070 | 26 | 30.6 (20.8 - 44.9) |
| | 20-24 years | 613210 | 206 | 33.6 (29.3 - 38.5) | 513674 | 170 | 33.1 (28.5 - 38.5) |
| | 25-29 years | 1419568 | 454 | 32.0 (29.2 - 35.1) | 1161363 | 374 | 32.2 (29.1 - 35.6) |
| | 30-34 years | 1437848 | 451 | 31.4 (28.6 - 34.4) | 1153007 | 366 | 31.7 (28.7 - 35.2) |
| | ≥ 35 years | 692805 | 205 | 29.6 (25.8 - 33.9) | 555688 | 172 | 31.0 (26.7 - 35.9) |
| Parity | 1st | 1744831 | 539 | 30.9 (28.4 - 33.6) | 1413085 | 449 | 31.8 (29.0 - 34.9) |
| | 2nd | 1550257 | 473 | 30.5 (27.9 - 33.4) | 1259698 | 380 | 30.2 (27.3 - 33.4) |
| | 3rd | 711277 | 242 | 34.0 (30.0 - 38.6) | 583463 | 201 | 34.4 (30.0 - 39.6) |
| | 4th | 186938 | 71 | 38.0 (30.1 - 47.9) | 154877 | 61 | 39.4 (30.6 - 50.6) |
| | ≥5th | 70350 | 22 | 31.3 (20.6 - 47.5) | 57684 | 17 | 29.5 (18.3 - 47.4) |
| Caesarean section | No | 3605606 | 1122 | 31.1 (29.3 - 33.0) | 2945515 | 927 | 31.5 (29.5 - 33.6) |
| | yes | 658046 | 225 | 34.2 (30.0 - 39.0) | 523292 | 181 | 34.6 (29.9 - 40.0) |
| Paternal age | <25 years | 339151 | 120 | 35.4 (29.6 - 42.3) | 284949 | 100 | 35.1 (28.8 - 42.7) |
| | 25-29 years | 1063260 | 331 | 31.1 (28.0 - 34.7) | 872111 | 287 | 32.9 (29.3 - 36.9) |
| | 30-34 years | 1516576 | 476 | 31.4 (28.7 - 34.3) | 1224784 | 378 | 30.9 (27.9 - 34.1) |
| | 35-39 years | 906455 | 288 | 31.8 (28.3 - 35.7) | 732697 | 237 | 32.3 (28.5 - 36.7) |
| | ≥40 years | 411728 | 126 | 30.6 (25.7 - 36.4) | 332815 | 100 | 30.0 (24.7 - 36.6) |
| **Maternal education** | | | | | | | |
| | Lower secondary | 582338 | 174 | 29.9 (25.8 - 34.7) | 489260 | 146 | 29.8 (25.4 - 35.1) |
| | Started upper secondary | 167607 | 54 | 32.2 (24.7 - 42.1) | 136935 | 43 | 31.4 (23.3 - 42.3) |
| | Completed upper secondary (high school or vocational education | 1226071 | 433 | 35.3 (32.1 - 38.8) | 1016683 | 364 | 35.8 (32.3 - 39.7) |
| | Post-secondary non-tertiary (started but not completed any higher degree) | 98838 | 35 | 35.4 (25.4 - 49.3) | 80765 | 27 | 33.4 (22.9 - 48.7) |
| | Bachelor's degree or equivalent | 1756924 | 547 | 31.1 (28.6 - 33.9) | 1415143 | 451 | 31.9 (29.1 - 35.0) |
| | Master's degree or equivalent | 398829 | 96 | 24.1 (19.7 - 29.4) | 304720 | 71 | 23.3 (18.5 - 29.4) |
| | PhD level | 28737 | 8 | 27.8 (13.9 - 55.7) | 21813 | 6 | 27.5 (12.4 - 61.2) |

*Information on maternal smoking was available for births from 1999. Providing information is optional and was provided by 81.4% of the person-years under observation. T1D: Type 1 diabetes. PYR: Person-years of observation.

**Table S4. Robustness analysis: Associations of maternal education with incidence of type 1 diabetes after further adjustment for maternal type 1 diabetes***

| Maternal education | | | | |
|---|---|---|---|---|
| | Period 1 (1989-2003) | Period 1 with additional adjustment for maternal T1D* | Period 2 (2005-2013) | Period 2 with additional adjustment for maternal T1D* |
| | aIRR (95% CI)† | aIRR (95% CI)† | aIRR (95% CI)† | aIRR (95% CI)† |
| Lower secondary | 1.04 (0.91 - 1.20) | 1.04 (0.91 - 1.20) | 0.94 (0.83 - 1.06) | 0.84 (0.70 - 1.00) |
| Started upper secondary | 0.93 (0.83 - 1.03) | 0.93 (0.83 - 1.03) | 0.89 (0.74 - 1.06) | 0.81 (0.61 - 1.07) |
| Completed upper secondary (high school or vocational education | 1 (Reference) | 1 (Reference) | 1 (Reference) | 1 (Reference) |
| Post-secondary non-tertiary (started but not completed any higher degree) | 1.03 (0.89 - 1.19) | 1.03 (0.89 - 1.19) | 0.95 (0.73 - 1.22) | 0.98 (0.69 - 1.38) |
| Bachelor's degree or equivalent | 0.86 (0.75 - 1.00) | 0.86 (0.75 - 0.99) | 0.90 (0.81 - 0.99) | 0.92 (0.81 - 1.05) |
| Master's degree or equivalent level | 0.79 (0.57 - 1.07) | 0.79 (0.58 - 1.08) | 0.83 (0.70 - 0.99) | 0.77 (0.61 - 0.97) |
| PhD level | 0.74 (0.10 - 5.27) | 0.73 (0.10 - 5.22) | 0.74 (0.41 - 1.34) | 0.87 (0.43 - 1.77) |

* Maternal type 1 diabetes was available with less than complete coverage for mothers in period 1 and from the Medical Birth Registry for births from 1999 onwards (Period 2).

† Incidence rate ratios (aIRR) for type 1 diabetes, adjusted for maternal age, parity, caesarean section, county of residence, and child's sex. CI: Confidence interval.

**Table S5. Robustness analysis: Associations of maternal and paternal education with childhood-onset type 1 diabetes if using parental education ascertained at different time-points***

| **Maternal education** | | | |
|---|---|---|---|
| | Period 1 (1989-2003): main analysis (education near birth of child) | Period 1: used subsequent available information on education if missing)* | Period 1: used education in the year 2001 for all subjects |
| | aIRR (95% CI)† | aIRR (95% CI)† | aIRR (95% CI)† |
| Lower secondary | 1.04 (0.91 - 1.20) | 1.04 (0.91 - 1.20) | 1.12 (0.96 - 1.30) |
| Started upper secondary | 0.93 (0.83 - 1.03) | 0.93 (0.84 - 1.04) | 0.96 (0.86 - 1.07) |
| Completed upper secondary (high school or vocational education | Ref. | Ref. | Ref. |
| Post-secondary non-tertiary (started but not completed any higher degree) | 1.03 (0.89 - 1.19) | 1.03 (0.89 - 1.20) | 1.28 (1.02 - 1.61) |
| Bachelor's degree or equivalent | 0.86 (0.75 - 1.00) | 0.86 (0.75 - 0.99) | 0.90 (0.79 - 1.01) |
| Master's degree or equivalent level | 0.79 (0.57 - 1.07) | 0.79 (0.58 - 1.07) | 0.87 (0.67 - 1.14) |
| PhD level | 0.74 (0.10 - 5.27) | 0.74 (0.10 - 5.25) | 0.43 (0.11 - 1.71) |
| **Paternal education** | | | |
| | Period 1 (1989-2003): main analysis (education near birth of child) | Period 1: used subsequent available information on education)* | Period 1: used education in the year 2001 for all subjects |
| | aIRR (95% CI)† | aIRR (95% CI)† | aIRR (95% CI)† |
| Lower secondary | 0.86 (0.75 - 0.98) | 0.86 (0.75 - 0.98) | 0.85 (0.73 - 0.99) |
| Started upper secondary | 0.97 (0.87 - 1.08) | 0.97 (0.87 - 1.08) | 1.05 (0.94 - 1.16) |
| Completed upper secondary (high school or vocational education | Ref. | Ref. | Ref. |
| Post-secondary non-tertiary (started but not completed any higher degree) | 0.88 (0.76 - 1.03) | 0.88 (0.76 - 1.03) | 0.86 (0.69 - 1.07) |
| Bachelor's degree or equivalent | 0.85 (0.72 - 1.00) | 0.86 (0.73 - 1.01) | 0.93 (0.82 - 1.05) |
| Master's degree or equivalent level | 0.96 (0.80 - 1.15) | 0.97 (0.81 - 1.16) | 0.95 (0.80 - 1.13) |
| PhD level | 1.11 (0.53 - 2.34) | 1.11 (0.52 - 2.33) | 0.86 (0.51 - 1.47) |

* If education was missing in the survey closest to the birth of the child, we used the next available information, typically 10 years later.

† Incidence rate ratios (aIRR) for type 1 diabetes, adjusted for maternal age, parity, caesarean section, county of residence, and child's sex. CI: Confidence interval.

**Table S6. Characteristics of all subjects in the first period (1989-2003) and the subgroup with information on parental occupations**

| Covariate | | All subjects in period 1989-2003 | | | Subjects in period 1989-2003 with information on parental occupation | | |
|---|---|---|---|---|---|---|---|
| | | T1D cases | Person-years | Incidence rate per 100 000 PYR (95% CI) | T1D cases | Person-years | Incidence rate per 100 000 PYR (95% CI) |
| | Total | 2325 | 9192165 | 25.3 (24.3 - 26.3) | 1497 | 5558402 | 26.9 (25.6 - 28.3) |
| Sex | Boys | 1242 | 4723688 | 26.3 (24.9 - 27.8) | 802 | 2853749 | 28.1 (26.2 - 30.1) |
| | Girls | 1083 | 4468300 | 24.2 (22.8 - 25.7) | 695 | 2704521 | 25.7 (23.9 - 27.7) |
| Maternal age at delivery | <20 y | 79 | 269577 | 29.3 (23.5 - 36.5) | 32 | 103119 | 31.0 (21.9 - 43.9) |
| | 20-24 y | 544 | 2261783 | 24.1 (22.1 - 26.2) | 325 | 1333472 | 24.4 (21.9 - 27.2) |
| | 25-29 y | 915 | 3661699 | 25.0 (23.4 - 26.7) | 647 | 2412386 | 26.8 (24.8 - 29.0) |
| | 30-34 y | 589 | 2272315 | 25.9 (23.9 - 28.1) | 383 | 1350477 | 28.4 (25.7 - 31.3) |
| | ≥ 35 y | 198 | 726791 | 27.2 (23.7 - 31.3) | 110 | 358948 | 30.6 (25.4 - 36.9) |
| Parity | 1st | 990 | 3889582 | 25.5 (23.9 - 27.1) | 605 | 2224674 | 27.2 (25.1 - 29.5) |
| | 2nd | 870 | 3419412 | 25.4 (23.8 - 27.2) | 579 | 2158833 | 26.8 (24.7 - 29.1) |
| | 3rd | 348 | 1456259 | 23.9 (21.5 - 26.5) | 238 | 916999 | 26.0 (22.9 - 29.5) |
| | 4th | 93 | 329224 | 28.2 (23.1 - 34.6) | 58 | 202547 | 28.6 (22.1 - 37.0) |
| | ≥5th | 24 | 97688 | 24.6 (16.5 - 36.7) | 17 | 55349 | 30.7 (19.1 - 49.4) |
| Caesarean section | No | 2041 | 8146319 | 25.1 (24.0 - 26.2) | 1323 | 4948219 | 26.7 (25.3 - 28.2) |
| | yes | 284 | 1045846 | 27.2 (24.2 - 30.5) | 174 | 610183 | 28.5 (24.6 - 33.1) |
| Paternal age | <25 y | 315 | 1192733 | 26.4 (23.6 - 29.5) | 148 | 526070 | 28.1 (23.9 - 33.1) |
| | 25-29 y | 810 | 3288251 | 24.6 (23.0 - 26.4) | 562 | 2194065 | 25.6 (23.6 - 27.8) |
| | 30-34 y | 780 | 3046489 | 25.6 (23.9 - 27.5) | 546 | 1957218 | 27.9 (25.7 - 30.3) |
| | 35-39 y | 348 | 1313297 | 26.5 (23.9 - 29.4) | 212 | 736591 | 28.8 (25.2 - 32.9) |
| | ≥40 y | 72 | 351394 | 20.5 (16.3 - 25.8) | 29 | 144459 | 20.1 (14.0 - 28.9) |

PYR: Person-years of follow-up. T1D: Type 1 diabetes. CI: Confidence interval.

**Table S7. Explorative analyses: Subcategories of parental occupation and incidence of childhood-onset type 1 diabetes***

| **Maternal occupation** | **T1D cases** | **Incidence rate per 100 000 person-years (95% CI)** | **Incidence rate ratio (95% CI)** |
|---|---|---|---|
| 04: Nurse and assistant nurse | 186 | 30.2 (26.12 - 34.82) | 1 (Reference) |
| 06: Teaching professionals | 74 | 31.2 (24.83 - 39.17) | 1.03 (0.79 - 1.35) |
| 20: Office workers | 23 | 22.8 (15.14 - 34.28) | 0.76 (0.49 - 1.16) |
| 21: Office workers / stenographers | 23 | 27.4 (18.19 - 41.20) | 0.91 (0.59 - 1.40) |
| 29: Other clerical workers | 148 | 28.6 (24.38 - 33.64) | 0.95 (0.77 - 1.18) |
| 33: Trade work | 145 | 31.8 (27.01 - 37.41) | 1.05 (0.85 - 1.31) |
| 41: Farmers | 28 | 32.6 (22.53 - 47.26) | 1.08 (0.73 - 1.61) |
| 67: Postal workers | 33 | 31.8 (22.64 - 44.79) | 1.06 (0.73 - 1.53) |
| 71: Tailors, sewers and related trades workers | 20 | 36.5 (23.56 - 56.60) | 1.21 (0.76 - 1.92) |
| 91: Hotel, restaurant and household workers | 60 | 18.3 (14.23 - 23.60) | 0.61 (0.45 - 0.81) |
| 92: Serving work | 21 | 21.0 (13.67 - 32.16) | 0.70 (0.44 - 1.09) |
| 93: Cleaners | 43 | 23.5 (17.41 - 31.66) | 0.78 (0.56 - 1.08) |
| 0X: Miscellaneous other professionals | 25 | 34.6 (23.38 - 51.20) | 1.15 (0.76 - 1.74) |
| **Paternal occupation** | **T1D Cases** | **Incidence rate per 100 000 person-years (95% CI)** | **Incidence rate ratio (95% CI)** |
| 00: Technical, scientific, humanistic, and artistic work | 52 | 20.7 (15.81 - 27.22) | 0.76 (0.56 - 1.05) |
| 06: Teaching professionals | 49 | 29.6 (22.34 - 39.11) | 1.09 (0.79 - 1.50) |
| 11: Corporate and organizational management | 26 | 29.2 (19.88 - 42.89) | 1.08 (0.71 - 1.63) |
| 29: Other clerical workers | 44 | 33.2 (24.68 - 44.56) | 1.22 (0.87 - 1.71) |
| 33: Trade workers | 78 | 32.1 (25.73 - 40.11) | 1.18 (0.90 - 1.56) |
| 40: Farmers | 39 | 26.1 (19.06 - 35.70) | 0.96 (0.68 - 1.37) |
| 41: Farmer workers | 42 | 28.0 (20.69 - 37.88) | 1.03 (0.73 - 1.45) |
| 64: Drivers | 83 | 27.8 (22.38 - 34.41) | 1.02 (0.78 - 1.34) |
| 75: Metal, machinery, and related trade workers | 153 | 27.1 (23.15 - 31.78) | 1 (Reference) |
| 76: Electrical and electronic trades workers | 79 | 28.9 (23.16 - 35.99) | 1.06 (0.81 - 1.40) |
| 77: Woodworkers (e.g. carpenters and sawmill workers) | 94 | 28.2 (23.03 - 34.50) | 1.04 (0.80 - 1.34) |
| 79: Other building and construction work | 28 | 20.7 (14.30 - 29.99) | 0.76 (0.51 - 1.14) |
| 82: Food processing and related trades workers | 28 | 26.5 (18.31 - 38.41) | 0.98 (0.65 - 1.46) |
| 85: Other manufacturing worker | 20 | 44.3 (28.55 - 68.58) | 1.63 (1.02 - 2.60) |
| 87: Construction machine drivers | 42 | 28.1 (20.74 - 37.97) | 1.03 (0.74 - 1.46) |
| 88: Storing and goods handling labourers | 32 | 24.9 (17.59 - 35.17) | 0.92 (0.63 - 1.34) |
| 91: Hotel, restaurant, and household worker | 25 | 42.8 (28.94 - 63.37) | 1.58 (1.03 - 2.41) |
| 0X: Miscellaneous other professionals | 31 | 33.4 (23.47 - 47.46) | 1.23 (0.84 - 1.81) |
| A1: Armed forces | 63 | 25.5 (19.89 - 32.59) | 0.94 (0.70 - 1.26) |

* The next highest level of detail in the Nordic occupational category coding (2 digits) included 82 categories. The table shows results for the 13 maternal occupation categories (representing a total of 829 T1D cases) and 19 paternal occupation categories (representing a total of 1008 T1D cases) which satisfied the predefined inclusion criterion of having at least 20 observed cases of incident type 1 diabetes. The category with the highest number of T1D cases were chosen as the reference category separately for maternal and paternal occupations. Codes according to the Nordic Classification of Occupations. T1D: Type 1 diabetes. CI: Confidence Interval. https://www.nb.no/nbsok/nb/8d2a147fbb144dbb1fa40670315cb966?lang=no#0

### Supplementary results: Figures

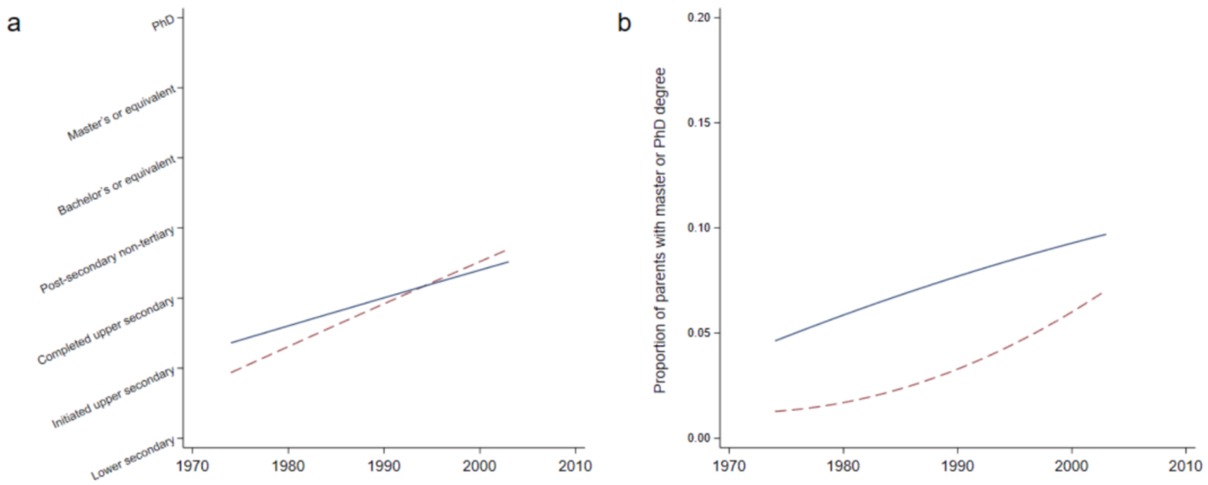

**Figure S1. Time trends in parental education in Norway**
a) Mean parental education at child's birth in the period 1989-2003. b) Proportion of parents with master or PhD degree at the time of child's birth. Maternal education red dash line and paternal education in solid blue. Correlation between maternal and paternal education: Spearman coefficient=0.43.

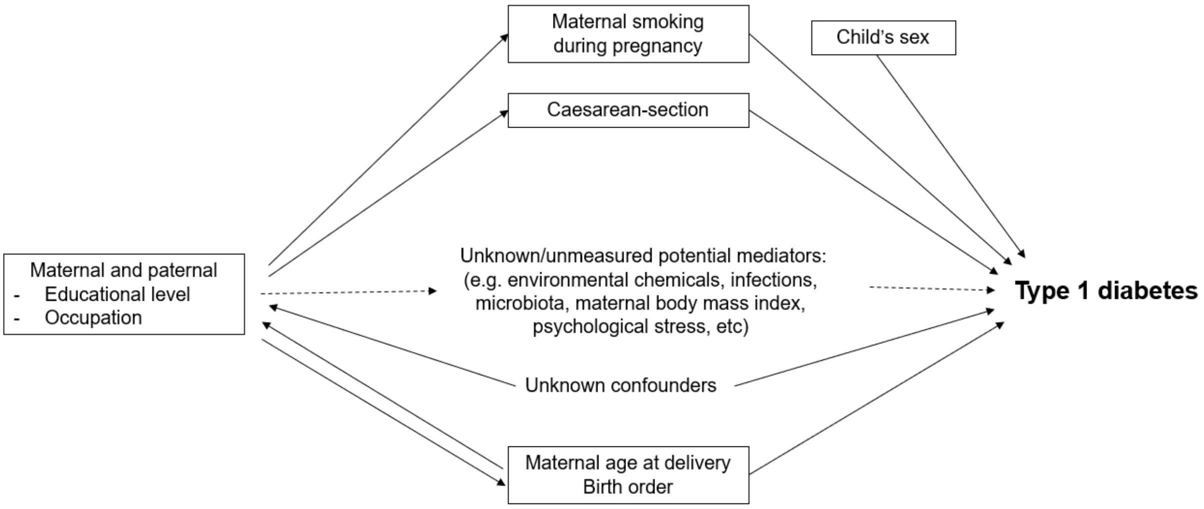

**Figure S2. Simplified causal model used as basis for interpreting analyses**
A conceptual model based on a priori knowledge and assumptions with necessary simplifications. Arrows represent assumed causal effects. Variables in solid boxes are adjusted for in our regression models. Maternal smoking was adjusted for in robustness analyses because it was only available in a subset. Although not shown in the figure, we also adjusted for calendar period, age and county of residence at birth in all models as explained in the methods section of the main text (and maternal type 1 diabetes in robustness analysis (Supplementary Table S4).

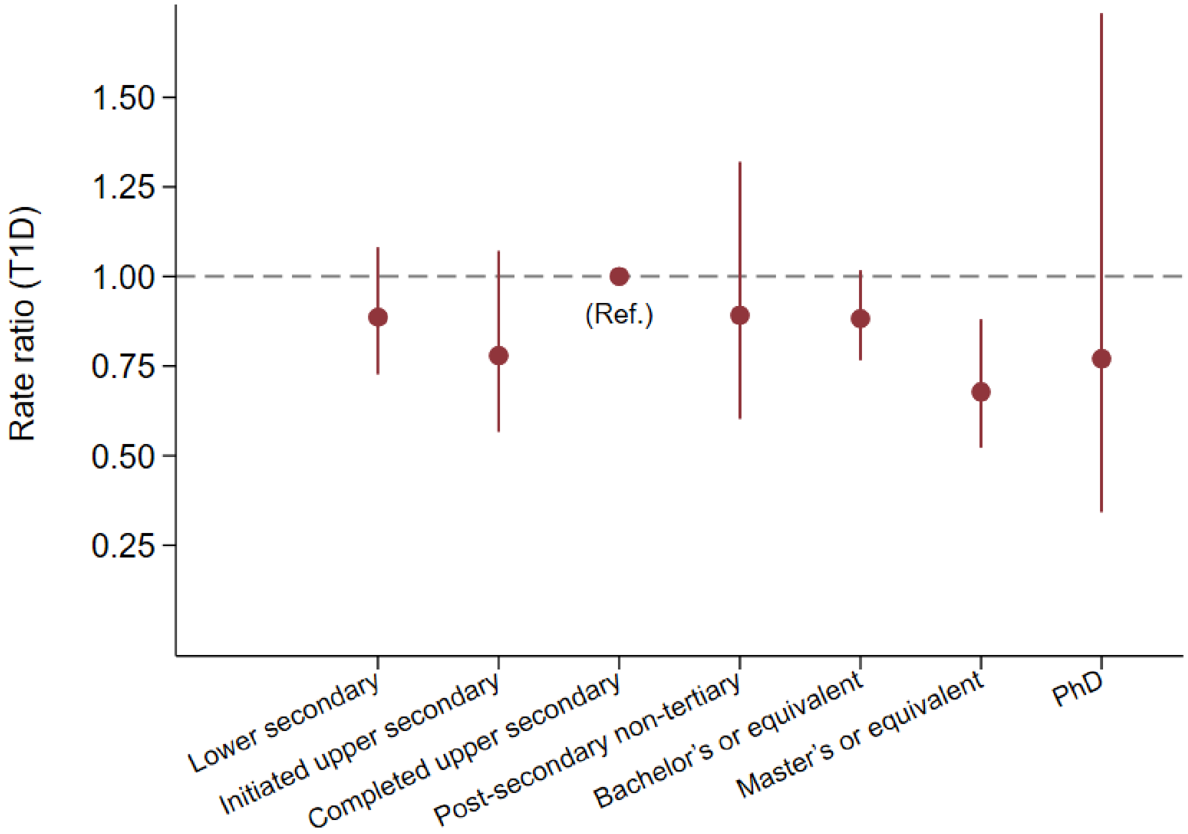

**Figure S3. Influence of adjustment for maternal smoking in the analysis of maternal education and risk of type 1 diabetes**
Information on maternal smoking was available for 81% of the births 1999-2013 (see detail characteristics in supplementary table S3). Incidence rate ratios adjusted for paternal age, parity, county of residence, child's sex and smoking. T1D: Type 1 diabetes.

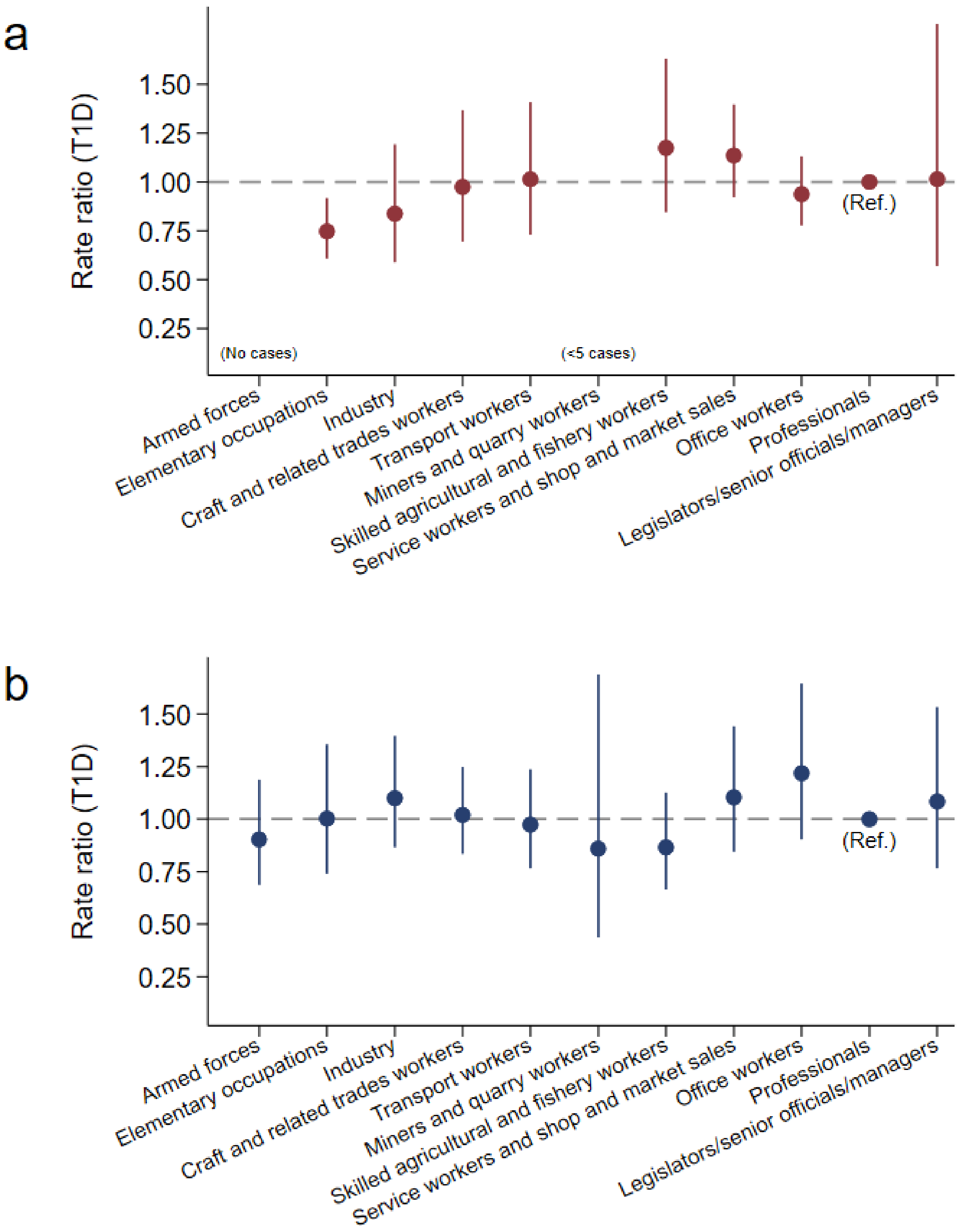

**Figure S4. Influence of additional adjustment for education in the analysis of parental occupation and risk of type 1 diabetes**

Likelihood ratio test (10 degree of freedom) p=0.014 for maternal occupation, likelihood ratio test=0.63 for paternal occupation. T1D: Type 1 diabetes.
